# Development and validation of a multiplexed quantitative PCR assay for clinical detection and surveillance of Oropouche virus

**DOI:** 10.64898/2026.05.26.26354109

**Authors:** Elyse Stachler, Kyle McMahon, Nisha Gopal, Hannah Knoll, Keith R. Baillargeon, Andrea C. Mora, Hunter A. Wondrash, Evan M. Sullivan, Sarah Rush, Dawn Gratalo, Al Ozonoff, Pardis Sabeti, Michael Springer

## Abstract

**Background:** Oropouche virus (OROV) is an emerging vector-borne virus with rapidly expanding geographic range, increasing case counts, and growing evidence of severe outcomes including neuroinvasive disease and vertical transmission. Because OROV infection presents with nonspecific febrile illness that overlaps clinically with other viruses including dengue, zika, and chikungunya, accurate molecular diagnostics are essential for patient care and surveillance. Yet existing assays rely on single genomic targets and are vulnerable to detection failure as the virus evolves and reassorts.

**Methodology/Principal Findings:** To support diagnostic capacity, we developed and clinically validated a multiplexed qPCR assay targeting three regions of the OROV S segment, incorporating redundancy to preserve sensitivity across viral diversity while enabling robust clinical interpretation. The multiplex also includes an assay targeting RNaseP as an internal sample control to ensure adequate sample processing. We evaluated assay performance using both historical and contemporary OROV strains and validated the assay on contrived serum, plasma, and cerebrospinal fluid samples, assessing linearity, limit of detection (LOD), accuracy, specificity, precision, and sample stability. The assay met or exceeded all predefined acceptance criteria for clinical testing and achieved an LOD as low as 6 copies per reaction for contemporary outbreak strains. We further implemented a logic-based interpretation matrix that reduced false-positive risk while maintaining sensitivity near the analytical LOD.

**Conclusions/Significance:** Our assay sensitively and specifically detects OROV RNA in serum, plasma, and cerebrospinal fluid while incorporating safeguards against viral evolution and reassortment. The assay has been approved for use by CLIA at Nexus Medical Labs in 49 U.S. states, expanding access to timely OROV diagnostics in the United States and providing a durable framework for molecular detection of reassorting, rapidly evolving viruses as OROV continues to spread into new regions.

**Author Summary:** We present a fully validated multiplex qPCR assay for detection of Oropouche virus (OROV) in clinical samples. Our multiplex design includes three assays targeting distinct regions of the S segment, protecting against assay failure due to mutation dropout. In addition, we include a human internal control assay (RnaseP), which gives confidence to pathogen-negative clinical results by ensuring sample quality and adequate sample processing. The assay exhibits high sensitivity and specificity, and can be adopted by other labs should expanded OROV surveillance be needed. With geographic and case number expansion of OROV in recent years, this assay adds to the available tools for surveillance and clinical diagnosis, mitigating negative effects of the virus in the future.

## Introduction

Oropouche virus (OROV) is a vector-borne virus endemic to regions in Central and South America and the Caribbean, where an outbreak in 2023-2024 resulted in nearly 60 times more cases than reported in previous years(1). The outbreak included cases in several non-endemic areas and travel-associated cases in countries that had not previously reported OROV, including the United States. Although the risk of OROV in the U.S. remains low(2), increased geographic spread has raised concerns(3), highlighting the need for expanded research, surveillance, and access to clinical diagnostics(4,5).

OROV causes Oropouche disease, a typically mild, self-limited illness characterized by fever, headache, and myalgia(6), but recent outbreaks have reported increasing disease severity(1) including neuroinvasive disease and capacity for vertical transmission between mother and fetus, causing birth defects and miscarriage(5,7,8). These symptoms overlap with other common arboviral infections in the region such as dengue, chikungunya, and zika, contributing to under-detection of OROV cases. Together, nonspecific clinical presentation and emerging evidence of more severe outcomes underscores the need for improved access to validated diagnostics to support accurate detection, inform patient care, and enhance surveillance(9–11).

OROV is a negative-sense, single-stranded RNA virus of the genus *Orthobunyavirus*, with a segmented genome that readily undergoes reassortment. To date, researchers have identified three OROV reassortants — Madre de Dios virus, Iquitos virus, and Perdões virus — which share the S and L genome segments but carry distinct M segments(12). Genomic analyses have shown the 2023-2024 outbreak was driven by a previously unrecognized OROV reassortant that likely emerged approximately a decade ago(13–15). This strain shows increased replication efficiency and virulence compared with historical strains(1), and has been associated with neuroinvasive disease and adverse pregnancy outcomes, highlighting how ongoing viral evolution can alter disease dynamics and complicate molecular detection due to sequence mutation and assay dropout (5,7,8,16).

In the United States, only the Centers for Disease Control and Prevention (CDC) performed diagnostic testing for suspected OROV cases at the time of the project start, substantially limiting access to testing should new cases or outbreaks occur. More broadly, existing molecular assays for OROV rely on single genomic targets, creating vulnerability to detection failure as the virus evolves and reassorts(17–19). Reliable clinical testing also requires internal controls to verify sample quality and nucleic acid extraction across diverse specimen types. Together, these constraints highlight the need for diagnostic assays that combine redundancy across viral targets with rigorous clinical validation and interpretable results across relevant sample matrices(4,10,11,14).

In response to a CDC call to rapidly expand diagnostic capacity, we developed and clinically validated a robust multiplexed qPCR assay for OROV detection within 60 days of project initiation. The assay targets three distinct OROV regions of the viral S segment, including two previously described assays and one newly designed assay to improve sensitivity and reduce susceptibility to viral evolution and reassortment. We combine these with an internal human control target (RNaseP), to ensure adequate clinical sample collection and processing. We evaluated assay performance using historical and contemporary OROV strains and validated the assay on serum, plasma, and cerebrospinal fluid samples, assessing limit of detection (LOD), linearity, sensitivity, specificity, precision, and sample stability. The assay met all predefined validation criteria and has been approved for use by CLIA at Nexus medical labs in 49 U.S. states, expanding access to OROV diagnostics and strengthening surveillance and preparedness as OROV continues to spread into new regions.

## Methods

### Molecular assay design and selection

For assay development, we evaluated 12 OROV singleplex candidates for selection into our multiplex panel, assessing nine novel designs alongside three previously designed assays. For the nine novel designs, we selected initial candidate amplicon sequences from conserved regions of the S and L segments of the OROV genome using ADAPT (Activity-informed Design with All-inclusive Patrolling of Targets)(20). Based on the ADAPT-identified amplicon regions, we designed candidate primers and probes for qPCR functionality using Integrated DNA Technology (IDT)’s Primer Quest tool, with optimal melting temperatures of 60°C and 70°C, respectively, as well as minimal secondary structure and dimerization potential. We aligned the resulting qPCR primer-probe sets to the OROV genome to confirm specificity using MAFFT v7.490 implemented in Geneious Prime (Biomatters Ltd.). Of the nine novel assays designed, two target the OROV S segment and seven target the OROV L segment. For positive controls, we generated synthetic gene fragments for each candidate assay by using S and L segment alignments as input for ADAPT’s target design function to identify representative target regions. Table S1 lists primer sequences for the nine novel assays and Table S2 provides positive control sequences.

For the three previously designed OROV assays, we assessed designs that all target the S segment: Naveca et al’s OROV assay which is currently used by CDC (S_CDC)(19), IDT’s OROV assay which is almost identical to CDC’s assay design but features a shorter locked nucleic acid probe (S_IDT), and Rojas et. al’s OROV assay which features a variable nucleotide in the probe (S_Rojas)(17). To remove overlap in genome coverage between the S_IDT and S_Rojas OROV assay designs, we designed a new forward primer for the S_Rojas assay using Geneious Prime 2025.0.1 (S_Rojas_mod). Lastly, following OROV assay multiplexing, we included the CDC’s previously designed RNaseP assay (RP_CDC) to act as an internal assay control. RP_CDC has previously been used in diagnostic tests with FDA approval for detection of SARS-CoV-2 in clinical samples. Table S3 lists primer sequences of published assays used in this study and Table S4 provides positive control sequences

### *In silico* analysis of designs

We evaluated primers and probes for potential cross-reaction with unintended targets using NCBI BLAST. In addition, CDC provided a list of pathogens (Table S10) whose genomes were downloaded from GenBank (accessed November 2024) and aligned with candidate primers and probes in Geneious, allowing for up to two mismatches per primer or probe. We analyzed *in silico* sensitivity by mapping the OROV assays against OROV S segment sequences >700bp obtained from GenBank (accessed October 2025) and aligned in Geneious, representing sequences collected from 1955-2024.

### Samples and controls

We used synthetic gene fragments (Twist Biosciences and IDT) of the OROV S segment and RNaseP gene as positive controls for qPCR standard curves. For sample matrix experiments, we spiked OROV RNA into three sample matrices – human serum and plasma (Research Blood Components, Watertown, MA) and synthetic CSF (SeraCare, 0175-0007) – to simulate patient samples. US CDC provided standard OROV genomic material, including a contemporary strain from the 2023-2024 outbreak (strain 240023, 2024, Cuba). Dr. Sean P. J. Whelan’s lab at Washington University, St. Louis, USA provided OROV strains R10 and R5 (1957 prototype strain TRVL 9760). As well, CDC provided Fast Technology Analysis (FTA) cards containing RNA from closely related viral species (Dengue, Zika, West Nile, Mayaro, and Chikungunya viruses). In addition to collaborator-sourced materials, we utilized viral stocks of Zika (ATCC VR-1528) and Dengue (ATCC VR-1856) viruses along with two control panels from Zeptometrix (Meningitis/Encephalitis panel NATMEP-BIO and Blood Culture Identification 2 panel NATBCP2) for specificity testing. Table S5 lists all samples used for validation testing.

### Nucleic Acid Extraction and Purification

We performed automated RNA extraction using a KingFisher™ Flex Magnetic Particle Processor (ThermoFisher) with 96 Deep-Well Head and MagMAX CORE Nucleic Acid Purification Kit (ThermoFisher) following the “Simple Workflow” with 100μL of sample input. A batch can run up to 94 patient test samples and two process controls (one positive and one negative control).

### Initial qPCR testing and assay down-selection

To down-select newly designed assays prior to multiplexing, we first ordered and tested all designs as singleplex FAM assays with primer and probe concentrations of 400 and 200nM, respectively. We conducted all qPCR experiments on either a QuantStudio 6 Flex (Broad Institute) or QuantStudio 7 (Nexus Medical Labs) using the Luna Probe One-Step RT-qPCR Kit (No ROX, NEB, E3007E) run as 10μL reactions, following the manufacturer’s recommendation for reagent composition and thermocycling protocol. We generated standard curves using serial dilutions of synthetic gene fragments from 1E7 to 1E1 copies/reaction. We determined assay performance and down-selection based on sensitivity (ensuring designs met or exceeded a threshold of 10 copies/uL in-well or 100 copies/reaction) and qPCR standard curve efficiency (90-110%) as well as *in silico* specificity analysis, and validation on serial dilutions of the R5 and R10 viral stock RNA.

### Multiplex assay validation

We combined the top three performing OROV assay candidates with the CDC’s RNaseP assay to create a multiplex qPCR assay. We titrated final primer and probe concentrations by first keeping primer concentration for all assays constant at 200nM and varying the probe concentration (100nM, 150nM, 200nM, 250nM, 350nM, and 500nM). Next, we held the probe concentration for all assays constant at 150nM while the primer concentrations were varied (100nM, 200nM, 300nM, 400nM and 500nM). We selected final primer and probe concentrations to optimize the fluorescent signal and consistent detection across replicates. We also tested the final assay configuration in triplicate on viral R5 stock extract diluted in extracted plasma to ensure optimal performance.

We evaluated the finalized multiplex qPCR assay for linearity, limit of detection, accuracy, precision, specificity, and sample stability.

### Linearity

We conducted linearity studies using contrived samples of heat-inactivated viral R5 stock preparation in each matrix (serum, plasma, and synthetic CSF) at dilutions of known concentration, testing 10 replicates each at five dilutions of the target concentrations (1, 10, 100, 1000, and 10000 copies/uL) along with 10 negative replicates. Acceptance criteria is defined as all standard curves having a coefficient of determination (R^2^) >= 0.98.

### Limit of detection (LOD)

We conducted LOD studies for OROV PCR assays using contrived samples of heat-inactivated viral R5 stock preparation in each matrix (serum, plasma, and synthetic CSF) at dilutions of known concentration. The LOD is defined as the lowest copy number detected in at least 95% of replicates, testing 20 replicates each across six dilutions (62, 31, 16, 8, 4, and 0 copies/reaction) in each of the three matrices. In addition, we evaluated sensitivity performance on a contemporary OROV strain (240023), testing five replicates each across six dilutions (200, 100, 50, 25, 12, and 6 copies/reaction). Acceptance criteria is defined as an LOD that meets or exceeds a threshold of 10 copies/uL in-well or 100 copies/reaction. Final LOD is defined as the lowest concentration that resulted in at least 19 positive replicates out of 20.

### Accuracy

We evaluated accuracy by comparing measured vs. expected recovery of standard genomic material using contrived samples. We prepared contrived positive samples using contrived samples of heat-inactivated viral R5 stock preparation in each matrix (serum, plasma, and synthetic CSF) at known concentration, targeting 2x (n = 4), 4x (n = 7), 6x (n = 2), 8x (n = 2), 40x (n=5), and 400x LOD (n=10). We evaluated a total of 120 samples: 90 contrived positives and 30 negatives. For each matrix type (plasma, serum, and CSF) we tested 30 positive and 10 negative samples. Sensitivity is defined as the Positive Percent agreement (PPA), calculated as the percentage of known positive contrived samples correctly identified as positive. Specificity is defined as the Negative Percent Agreement (NPA), calculated as the percentage of known negative samples correctly identified as negative. Acceptance criteria is defined as PPA >= 95%, NPA >= 95%, and overall agreement >= 95%.

### Specificity

We evaluated molecular interference using FTA cards from the CDC of several closely related viruses, such as Dengue, Zika, West Nile, and Chikungunya (Table S5) as well as two panels from Zeptometrix (n=48). In total, we tested 58 samples for potential cross-reaction. We eluted nucleic acid from FTA cards by adding 100µL of phosphate buffered saline (PBS) to each 4-mm diameter FTA card punch. We extracted the FTA eluate and liquid viral stocks of Dengue and Zika (Springer Lab) following the protocol listed in section “**Nucleic Acid Extraction and Purification”** and subsequently ran triplicate extracts at stock concentration and 1:1000 dilutions (prepared post-extraction), while we tested Zeptometrix samples once each. Acceptance criteria is defined as no positive result returned for any of these off-target samples.

### Precision

We evaluated precision by testing the same standard material across two independent operators, using two instruments, across three days. We evaluated reproducibility using contrived samples of heat-inactivated viral R5 stock preparation in each matrix at known concentration with a target of 2,000x LOD. We made initial stocks at 31,260 copies/µL for serum and CSF and 15,620 copies/µL for plasma. We prepared each sample of spiked matrix on day one, aliquoted, and froze separately to ensure no freeze-thaw cycles over the three days of testing. Two operators tested all samples in triplicate on two sets of instruments (KingFisher Flex 1, KingFisher Flex 2, QuantStudio 1, and QuantStudio 2) across three days. Intra-run variability is determined by calculating the mean Ct value for all replicates for a given sample within a single OROV RT-PCR run. Inter-run variability is evaluated between runs performed by the same operator on different extraction and real-time PCR instruments and between operators. Acceptance criteria is defined as intra- and inter-run coefficient of variation (CV) <= 5%.

### Sample Stability

We evaluated sample stability by testing contrived positive samples (serum, plasma, and synthetic CSF) stored at cold (4° C) and frozen (-20° C) temperatures for 7 and 21 days for degradation. We prepared contrived positive samples using heat-inactivated OROV spiked into each matrix at known copies/µL with targets of 2x LoD (16 copies/reaction, n = 30), 3-5x LoD (31 copies/reaction, n = 10), and negatives (n=20). Acceptance criteria is defined as PPA >= 95%, NPA >= 95%, and overall agreement >= 95%.

### Clinical sample result interpretation

We established the interpretation matrix for the OROV multiplex assay during assay validation to define result classification thresholds. We analyzed a total of 798 negative clinical specimens (defined as matrix samples that have not been spiked with any viral material) using the QuantStudio platform with automatic thresholding to characterize background amplification. For any assay targets showing amplification in negative specimens, the preliminary cutoff is defined as the mean Ct of amplifying negatives minus three times the standard deviation (mean - 3xSD). We then refined these statistical thresholds using analytical sensitivity data at and near the LOD to ensure appropriate discrimination between low-level true positives and background signals. The final cutoff and indeterminate ranges are selected to maintain sensitivity at the LOD while minimizing false-positive interpretation.

### Clinical sampling readiness

We submitted the validated assays to the Massachusetts Department of Health which allows Nexus Medical Labs to support clinical operations in 49 states where Nexus holds relevant licensure. We also submitted the validation report to the Clinical Laboratory Evaluation Program division of the New York State for approval and licensure of the assay, which will upon approval allow Nexus Medical Labs to perform and report clinical results as well as serve as a reference laboratory for clinical sample processing in all 50 states until labs are able to validate, submit, and receive licensure to process their own samples.

## Results

### Development of a multiplex qPCR assay for OROV detection

To identify robust targets for multiplexed OROV detection, we designed nine novel qPCR assays targeting conserved regions of the S and L segments of the OROV genome (see methods for design details), and evaluated them alongside three previously reported S-segment assays (S_CDC, S_IDT, and S_Rojas_mod). In singleplex testing, the novel assays targeting the S segment achieved qPCR efficiencies of 90-102%, whereas assays targeting the L segment showed more variable efficiencies of 61-92%, with all designs exhibiting R^2^ values ≥ 0.95 (Figures S1-S2). The three previously described S-segment assays showed qPCR efficiencies between 88-110%, with R^2^ values ≥ 0.95 (Figure S3) when tested on viral standards.

Based on qPCR efficiency and sensitivity on positive control material, we selected three primer-probe sets targeting distinct regions of the OROV S segment (S_IDT, S_Rojas_mod, and S_G3; hereafter referred to as S1, S2, and S3) for inclusion in a multiplex assay. We configured the multiplex to detect S1 via SUN, while combining S2 and S3 for additive detection using FAM (referred to as S2/S3, R^2^ ≥ 0.98 for all matrices, See Figure 1) due to *in silico* mismatch analysis (see below). We also included an assay targeting the human RNaseP gene as an internal control to verify sample integrity and adequate nucleic acid extraction in OROV-negative clinical specimens. Table 1 summarizes the final primer-probe sequences, concentrations, fluorophore assignments, and assay source, while full assay composition and cycling conditions are provided in Tables S6 and S7, respectively.

**Figure 1:**
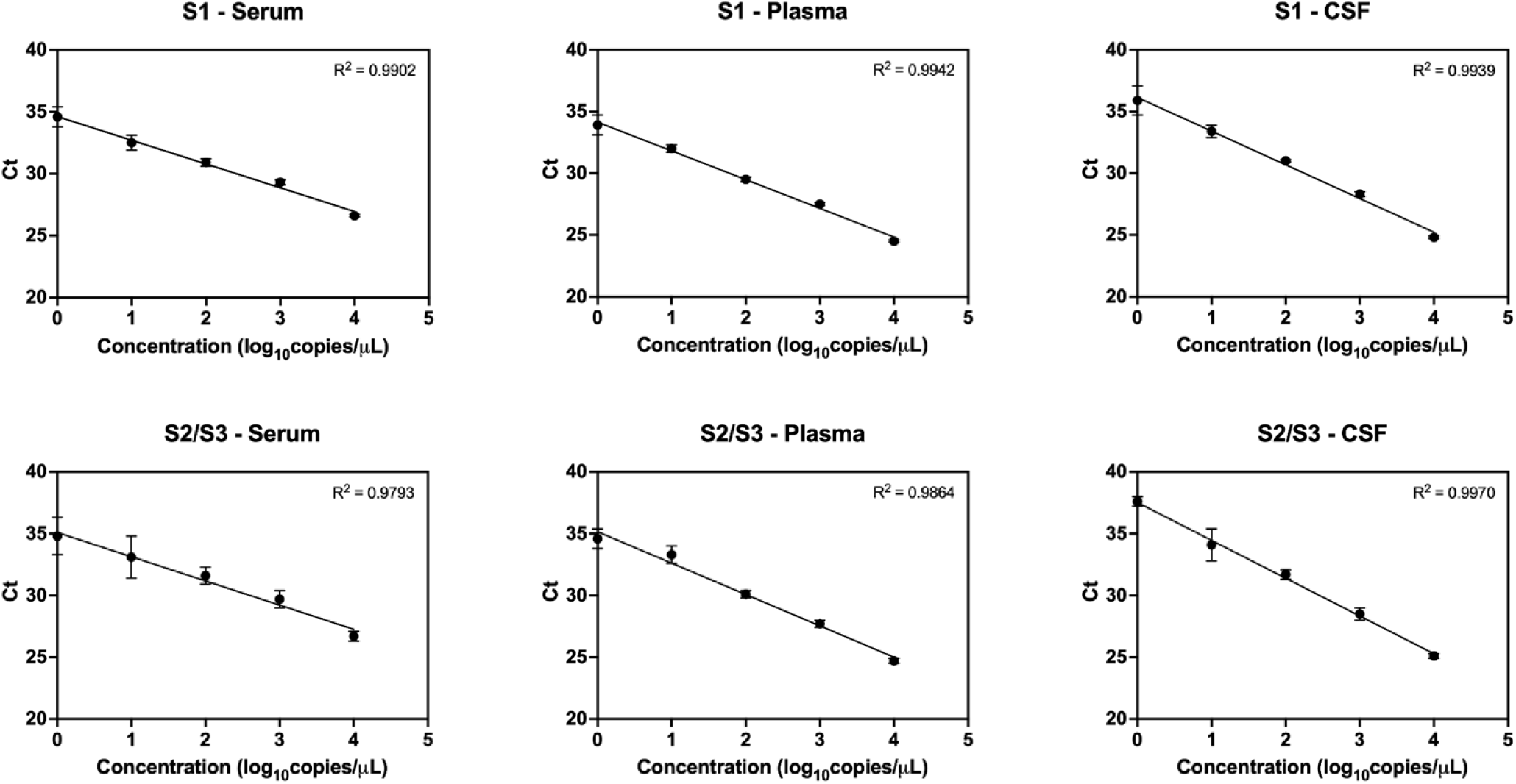
Linearity of the multiplex OROV qPCR assays across clinical sample matrices. We spiked viral RNA into serum, plasma, and cerebrospinal fluid (CSF). Data points represent the mean of 10 replicates, and error bars indicate ±1 standard deviation.

**Table 1:**
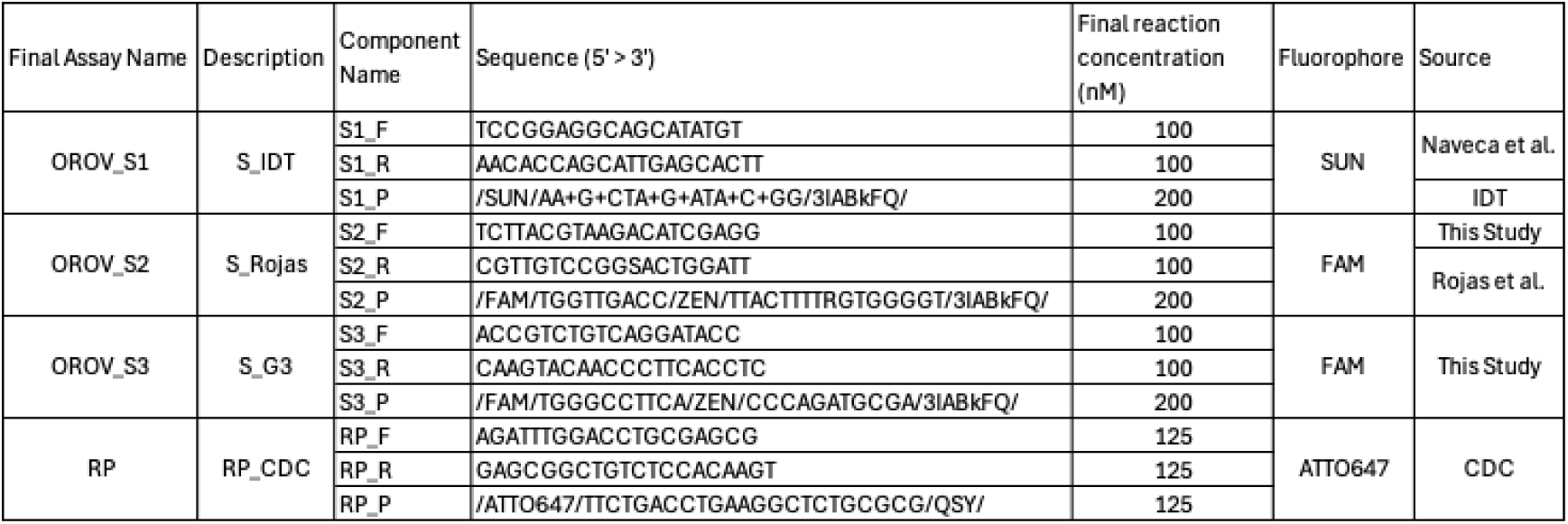
Multiplex OROV assay composition.

### *In silico* sensitivity and specificity analysis of multiplex assay designs

Since we performed wet-lab validation using both a contemporary OROV clinical isolate from 2024 (Strain 240023, Figure S3) and the prototype TRVL 9760 strain from 1957, we aligned the selected assays to these genomes to evaluate predicted detection (Figures S4 and S5). All primers matched the contemporary viral sequence exactly. In contrast, for the historical strain, S1 and S2 each contained a single mismatch in the forward primer, while S3 contained two mismatches in the probe and and three mismatches in the reverse primer.

To further assess *in silico* sensitivity of the selected OROV designs used in the multiplex assay, we mapped the individual primer-probe sets against 831 publicly available OROV S-segment sequences to estimate the percentage of circulating diversity predicted to be detected (see methods for details on alignments). The S1 and S2 assays aligned to 829 and 830 sequences (>99%), respectively, while the S3 assay aligned to 815 sequences (>98%). Of the 16 sequences that did not align to the S3 assay, 15 originated from samples collected more than 20 years ago, and all were collected more than 10 years ago, predating the estimated emergence of the current outbreak strain. Importantly, for any sequence where an assay was missing, there was at least one other assay predicted to detect the sequence. Together, these results indicate that the multiplex assay designs exhibit high coverage across known OROV sequence diversity while retaining sensitivity to contemporary circulating strains.

We assessed *in silico* specificity by mapping the selected OROV assays against genomes from other viral species, including distant relatives (e.g. flaviviruses and viruses from other genera), close relatives within the *Orthobunyavirus* genus, and known OROV reassortants (full list in Table S10). We observed no predicted cross-reactivity with either distant or close non-OROV viral relatives (allowing up to 2 mismatches in any primer or probe). Among OROV reassortants, the S1 assay aligned with 100% identity, the S2 assay aligned with one or two mismatches in the forward primer, and S3 showed 83-92% alignment (Figure S6). Together, these results indicate that the multiplex assay retains specificity for OROV, while differential alignment of the S3 assay may provide sensitivity to distinguish between reassortments.

### Analytical validation of the multiplex OROV qPCR assay

To validate our multiplex assay, we evaluated assay linearity, LOD, accuracy, precision, and sample stability using contrived clinical samples generated by spiking viral standard into serum, plasma, and cerebrospinal fluid (CSF). Table 2 summarizes assay performance, with all metrics meeting predefined acceptance criteria established to fulfill requirements for New York State Clinical Laboratory Evaluation Program (CLEP) requirements. Linearity studies showed that assay signal scaled directly with OROV concentration across all matrices, with high correlation (all R^2^ >= 0.98).

**Table 2:** Summary of analytical performance characteristics of the validated multiplex OROV qPCR assay.

| Validation Parameter | Acceptance criteria | Validation Results achieved | Pass/Fail |
| --- | --- | --- | --- |
| Linearity | $R^2 \geq 0.98$ | all curves $R^2 \geq 0.98$ | Pass |
| Limit of Detection (LOD) | Lowest concentration with 20/20 replicates detectable | LOD $\leq 12$ copies/reaction for S1 for all matrices.<br>LOD $\leq 6$ copies/reaction for S2/S3 for all matrices | Pass |
| Precision | Intra and Inter assay CV $\leq 5\%$ | Both Intra and Inter assay CVs $\leq 5\%$ | Pass |
| Accuracy | Over all Agreement $\leq 95\%$ Positive predictive agreement $\leq 95\%$ Negative predictive agreement $\leq 95\%$ | Overall Agreement $\geq 98.3\%$; PPA $\geq 96.6\%$; NPA = 100% | Pass |
| Molecular Interference and specificity | None of the closely related viruses, such as Dengue, Zika, West Nile, and Chikungunya organisms other than the Oropouche virus will be positive. | One false positive at high Ct (L. monocytogenes) listed as a limitation | Pass |
| Sample Stability | $\geq 95\%$ positive percent agreement $\geq 95\%$ negative percent agreement | Stable when stored at $-20^\circ\text{C}$ up to 21 days in all matrices | Pass |

The LOD for the contemporary OROV strain was 12 copies per reaction for S1 and 6 copies per reaction for S2/S3, whereas the prototype strain showed higher LODs of 31 and 62 copies per reaction for S1 and S2/S3, respectively, likely due to mismatches between the strain and primer and probe sequences. Accuracy testing yielded 100% positive predictive agreement (PPA) in serum, while one false negative was observed in each of plasma and synthetic CSF (PPA = 96.6%); both false negatives occurred at the lowest concentration tested (approximately 2xLOD). All 30 negative samples were correctly identified (negative percent agreement [NPA] =100%), resulting in an overall agreement of 100% for serum and 98.3% for plasma and CSF.

We assessed precision by testing the same standard material across two operators, two instruments, and three days. Both intra- and inter-run variability met the acceptance criteria, with coefficients of variation (CV) ≤ 5%. Intra-run CVs ranged from 0.6 - 1.3% for S1 and 0.7 - 1.7% for S2/S3 across all matrices. Inter-run CVs for S1 were 0.4%, 2.3%, and 4.7% for serum, plasma, and CSF, respectively, while S2/S3 showed inter-run CVs of 1.0%, 3.2%, and 4.0% for the same matrices.

We evaluated contrived sample stability over 21 days under refrigerated (4°C) and frozen (–20°C) storage conditions. Samples remained stable for at least 21 days in all three sample matrices at –20°C conditions and for at least seven days in serum and CSF when stored at 4°C.

In addition, we evaluated *in vitro* specificity by screening extracted nucleic acid from 58 viral and bacterial isolates (Table S5). Only one sample (*L. monocytogenes*) yielded a false positive result. The corresponding Ct value (36) fell within the predefined indeterminate range of the S1 assay and is therefore likely a false positive (see Interpretation matrix section below). Together, these results demonstrate that the assay provides sensitive and specific detection of OROV in clinical samples.

### Development of a decision matrix for clinical result interpretation

To ensure robust interpretation of clinical results, we developed an interpretation matrix that reduces false-positive detections while preserving assay sensitivity. We estimated the false-positive rates for each OROV target by evaluating Ct values from unspiked matrix controls during assay validation. The S1 assay showed background amplification in approximately 7.9% of 798 negative clinical specimens, with a mean Ct of 35.5 ± 0.9. This range overlapped with Ct values observed for low-copy true-positive samples (Ct of ∼35.2 at 2xLOD). In contrast, the OROV S2/S3 assay showed no false-positive amplification. Based on these findings, we applied initial Ct cutoffs of ≤ 35.0 for S1 and ≤ 36.0 for S2/S3 set above the average Ct observed at the LOD.

To address residual overlap between false-positive and true-positive signals near the assay LOD, we implemented a logic-based interpretation matrix (Tables S8 and S9). We interpret results that fall within defined Ct cutoffs and display valid amplification curves, characterized by a typical qPCR sigmoidal shape and clear exponential phase, as positive. We classify results within a predefined indeterminate zone (S1: 35.0 ≤ Ct ≤ 37.0; S2/S3: 36.0 ≤ Ct ≤ 38.0) as presumptive positives and subject them to repeat testing from the same extract. On repeat testing, we accept a slightly broader threshold (S1: Ct ≤ 37.0; S2/S3: Ct ≤ 38.0) to account for expected biological and technical variability at low-copy number. We report amplification that is not reproducible or lacks a valid curve as inconclusive or negative, based on internal control performance and predefined logic criteria. Together, this interpretation matrix preserves assay sensitivity while reducing, though not entirely eliminating, false-positive detections.

## Discussion

We developed and analytically validated a multiplexed qPCR assay for OROV that is CLIA approved for qualitative detection of viral RNA in serum, plasma, and CSF. The assay addresses a diagnostic need identified by the U.S. CDC by expanding access to OROV testing beyond centralized public health laboratories and by improving robustness to viral evolution. In contrast to prior OROV assays that rely on a single genomic target, our multi-amplicon design incorporates redundancy across the viral S segment together with an internal human control, strengthening confidence in both viral detection and upstream sample integrity.

Targeting the OROV S segment emerged as a central design principle. Consistent with prior studies, S-segment assays demonstrated strong analytical performance, but multiplexing revealed informative differences across targets. S1 and S2 aligned broadly across historical and reassortant OROV sequences, whereas S3 showed selective mismatches and enhanced sensitivity to contemporary outbreak strains. This pattern suggests that combining conserved and emergent targets can preserve detection across legacy diversity while maintaining sensitivity to newly circulating strains. In addition, differences in target amplification could alert to viral mutation and aid surveillance efforts, similar to patterns described in SARS-CoV-2 assay dropout that alerted to new mutations(21,22). Although differential alignment of S3 may offer sensitivity to reassortment, we interpret this as a potential analytical signal rather than a definitive discriminant feature, and additional validation across diverse clinical isolates will be required to assess its utility in distinguishing reassortant viruses.

Our analytical validation met all predefined acceptance criteria established by New York CLEP. The assay demonstrated strong linearity across matrices, low LODs for contemporary outbreak strains, and high accuracy and precision across operators, instruments, and days. As expected for assays operating near the LOD, we observed low-frequency background amplification for one target (S1), while the remaining targets showed no false positives. Rather than excluding a sensitive assay target, we addressed this tradeoff through a logic-based interpretation matrix that integrates Ct thresholds, amplification curve quality, internal controls, and repeat testing. This approach preserves sensitivity while reducing false-positive reporting and reflects the practical realities of clinical molecular testing for low-copy viral RNA.

Several limitations merit consideration. First, validation relied on a limited number of viral strains since no clinical samples were available to us. These strains spanned a wide temporal range but did not capture the full genetic diversity of circulating OROV. As clinical samples become available, further evaluation across patient-derived isolates and newly released OROV sequences will be important to confirm sustained performance. Second, while the S segment has been conserved across reassortants historically, it is possible that future strains could not follow this pattern. This does present a risk in our S-targeting strategy. In addition, although the interpretation matrix reduces false positives, it may introduce rare false-negative results near the assay LOD, a tradeoff inherent to highly sensitive molecular diagnostics. Finally, we observed one false-positive signal from a non-target organism (*L. monocytogenes*) that fell within the indeterminate range, underscoring the importance of curve-based interpretation and option to repeat testing of inconclusive results in clinical deployment.

A major strength of this work is the speed of translation from design to clinical authorization. In response to a CDC call to expand OROV diagnostic capacity, we developed, validated, and submitted this assay for clinical approval within 60 days. This timeline demonstrates that robust, regulatory-grade diagnostics for emerging pathogens can be designed and deployed rapidly when sequence data, assay design frameworks, and validation infrastructure are in place. The resulting assay provides clinicians with an additional option for diagnosing suspected OROV infection in the U.S. and can be adopted by other clinical laboratories or adapted for broader surveillance efforts.

OROV continues to pose a growing public health concern as its geographic range expands and recent outbreaks report increasing disease severity, including neuroinvasive disease and adverse pregnancy outcomes. Rapid and reliable molecular diagnostics are essential both for patient care and for surveillance as OROV spreads into new regions through travel and vector expansion. Beyond OROV, this work illustrates a generalizable approach to diagnostic design for reassorting, rapidly evolving arboviruses: multiplex targeting of conserved and variable regions, paired with rigorous analytical validation and transparent interpretation logic. Together, these principles can support durable diagnostic readiness for current and future emerging viral threats.

## Data Availability

All validation data is provided in the Supplemental Appendix.

## Acknowledgements

The authors would like to thank individuals from Ginkgo Bioworks (Birgitte Simen, Emily Schenkein, Mary Voneiff, Karen Hogan, Erin Turmelle, Peter Donaghue, Dan Bayley, Zachary Jacobson, and Tyler Clarkson), the Broad Institute (Bronwyn MacInnis), and Nexus Medical Labs (Esther Brown and Stephanie Meyer).

## Supporting Information Captions

**Table S1:** Primer and probe sequences for OROV and assays designed by the Broad Institute.

**Table S2:** Gene fragment sequences for OROV assays designed by the Broad Institute.

**Table S3:** Primer and probe sequences for predesigned OROV and RNaseP assays.

**Table S4:** Gene fragment sequences for OROV and RNaseP predesigned assays.

**Table S5:** Samples used for challenge experiments with source.

**Table S6:** Reaction composition for the final validated multiplex OROV assay.

**Table S7:** Thermocycling conditions for the final validated multiplex OROV assay.

**Table S8:** Interpretation matrix for initial run on clinical samples.

**Table S9:** Interpretation matrix for retesting on clinical samples after an inconclusive initial run.

**Table S10:** CDC provided list of pathogens whose genomes were downloaded from GenBank to screen for *in silico* specificity.

**Figure S1:** Results of Broad S segment assays on synthetic gene fragments.

**Table S11:** Assay characteristics of Broad S segment assays on synthetic gene fragments.

**Figure S2:** Results of Broad L segment assays on synthetic gene fragments.

**Table S12:** Assay characteristics of Broad L segment assays on synthetic gene fragments.

**Figure S3:** Results of pre-designed assays on viral seedstocks (R5 and R10).

**Figure S4:** Alignment of tested S segment assays to the OROV genome S segment originating from a 2024 Cuba Traveler (240023). The color of the primers indicates if it is a forward primer (dark green), reverse primer (light green), or probe (red).

**Figure S5:** Alignment of tested S segment assays to the OROV genome S segment originating from the 1957 prototype strain TRVL 9760. The color of the primers indicates if it is a forward primer (dark green), reverse primer (light green), or probe (red).

**Figure S6:** Alignment of tested S segment assays to the Perdoes virus genome S segment as an example of off-target specificity. The color of the primers indicates if it is a forward primer (dark green), reverse primer (light green), or probe (red).

